# Development and Performance verification of colloidal gold labeled SARS-CoV-2 antigen detection method for routine popular screening of COVID-19 with clinical samples in Poland and China

**DOI:** 10.1101/2021.11.22.21266719

**Authors:** Chuanxiang Guo, Li Yao, Fengling Chen, Chao Zhang, Wei Chen

**Author notes:** As we are continuing the further optimization of our detection methods and protocols, we will be uploading the updated results. Questions and feedback should be addressed to W. C.

## Abstract

In this research, we have constructed and optimized the colloidal gold labeled lateral flow strip (LFS) for rapid detection of antigen of SARS-CoV-2 and rapid screening of COVID-19. Based on the constructed and optimized colloidal gold lateral flow strip, the parameters of the LFS have been well evaluated with the clinical samples in the professional labs. The screening performance have also been evaluated from the aspects including the CT values, age distribution and onset of symptoms. Finally, based on the detection results of 420 clinical samples, the LFS can achieve the screening of COVID-19 with the positive percentage agreement (PPA, sensitivity), negative percent agreement (NPA, specificity), the positive predictive value (PPV) and the negative predictive value (NPV) of 96.8%, 100%, 100% and 96.6%, respectively, indicating the powerful potential for practical screening applications in pandemic control. Of great significance, this developed SARS-CoV-2 antigen detection method has also been successfully utilized for screening of delta-variant of SARS-CoV-2.

## A. Overview

The first outbreak of COVID-19 in Wuhan, China, 2019 and the subsequent prevalence of COVID-19 pandemic have caused serious life and economic loss all over the world ^1^. With the confirmed whole genomic sequence information, the current gold standard method for SARS-CoV-2 detection is still the real time reverse transcript quantitative PCR ^2^. With the evolution of the virus and the effective control of the pandemic in most of the countries, real time PCR can meet the detection requirements of acute outbreak in specific cities. However, with the resumption of work and production in many countries, routine screening of COVID-19 has become an effective strategy to prevent the outbreak of the pandemic. The large-scale screening in school, hospital, airport, station, and prison has been extensively conducted in Europe and US et al. Under this circumstance, the nucleic acid-based detection methods absolutely cannot the meet the urgent and huge requirements of common populations. The easy operation procedures and simple results judgement of colloidal gold labeled lateral flow strips make it suitable for wide applications in high throughput screening of common populations ^3-5^. Herein, we systematic optimized and screened lots of raw materials to construct the colloidal gold labeled COVID-19 antigen detection kit for rapid and easy detection of N protein of SARS-CoV-2. As we know that the coding gene of N protein is comparable conservative and the N protein is comparable stable, which determine that the developed colloidal gold labeled COVID-19 antigen detection kit can also be adopted for rapid screening of delta variants.

## B. Materials and Reagents

The antibody for gold nanoparticle labeling and antibody for coating on nitrocellulose membrane (NC membrane) as detect line were ordered from Chongqing Fantibdoy S&T Co. Ltd. The goat-anti-mouse was also ordered from the same provider. The NC membrane (ealon 120B) was purchased from the Ealon Membrane (Shantou, China) Co. Ltd. Other materials including the adhesive pad (300*60 mm), the conjugation pad (KB50) and the sample pad (8965) were bought from the Shanghai Jiening Biotechnol. Co. Ltd.

## C. Experimental Protocol

Firstly, the colloidal gold nanoparticles were prepared according to our previous reports. Then K2CO3 was used to modify the pH of gold nanoparticles to conjugate with the antibody against N protein of SRAS-CoV-2. Then, the colloidal gold labeled antibody conjugates were further blocked with BSA (0.5%, wt.%) and reacted for 30 min. The mixture was centrifuged at 10000 r/min for 40 min and the supernatant was discarded. While the precipitates were redispersed in the suspension buffer (0.1M Tris+20% sucrose+5.6% trehalose+1.5% BSA,pH=7.8), which was used for the preparation of conjugation pad at the spraying rate of 2 μL/cm and dried at 37 °C for 2 h.

For the preparation of test line (T line) and control line (C line), the antibody against the N protein of SARS-CoV-2 (1.0 mg/mL) and goat-anti-mouse antibody (1.5 mg/mL) were sprayed onto the NC membrane at the speed of 1.0 μL/cm. Finally, all components of the lateral flow strip were assembled to obtain the colloidal gold labeled COVID-19 antigen detection kit. The assembled and packaged colloidal gold labeled COVID-19 antigen detection kit were adopted for the screening of SARS-CoV-2 directly.

The pharyngeal and nasopharyngeal swabs were used to collect the samples and put into the lysis solution in extraction tube and rotate for 10 seconds. The swab was pressed against the inner wall of the extraction tube to release the antigen in the swab. Squeeze the extraction tube to remove the liquid as much as possible from the swab. The swab was disposed and packaged into the biohazard bags. Two drops of the extraction solution were added into the sample pore of the lateral flow strip directly and the screening results can be judged within 20 min. Of note, the negative results should be judged and reported after 20 min and the results judged more than 30 min were no longer valid.

## C. Assay Specifications

For the verification of the properties of the developed colloidal gold labeled SARS-CoV-2 antigen detection kit, all samples were detected prospectively. Each collected sample was measured by both the developed colloidal labeled SARS-CoV-2 antigen detection kit and the gold standard real time PCR test kit performed on the LightCycler 480 Real-Time PCR system (Roche) and Cobas z480 Analyzer (Roche). There are also some requirements about the samples for test. (1) for the enrolled subjects, all subjects should be the suspected patients (asymptomatic patients can also be considered for enrollment); (2) for the onset time of symptoms, all samples should be collected from the subjects of the onset of symptoms within 7 days. Meanwhile, the collected samples can be divided into different groups according to the onset time of symptoms including ⩽3 days, 4-7 days and >7 days. And the percentages of these three groups can be set as 40%, 40% and 20%, respectively. (3) the collected samples should also be enrolled according to the age distributions including <14 years, 14-24 years, 25-45 years, 46-65 years and ⩾65 years and the corresponding percentages of each group can be set as 10-20%, 15-20%, 15-20%, 20-25%, and 30-35%, respectively. If the sample could not meet the above requirements for enrollment, the sample can be excluded not for verification.

All the samples collected should also be measured with the real time PCR. Considering the consistency of the sensitivity and specificity can be influenced by the viral loading of the patients, the results of RT-PCR in the form of CT values should cover different ranges. Meanwhile, in order to verify the accuracy of the kit, some weak positive samples were artificially enrolled into the groups for the evaluation.

## D. Results and Discussion

For the verification research with both the developed colloidal gold labeled SARS-CoV-2 antigen detection kit and the real-time PCR method. All enrolled subjects were 420 and there were 220 positive samples (Ct<38) confirmed with the real-time PCR. Then, all the tested results with the colloidal gold labeled SARS-CoV-2 antigen detection kit were further analyzed and compared with those of real time PCR.

Firstly, all the enrolled subjects were tested with our constructed method and the results were analyzed compared to results of the real time PCR in the form of Ct values. From results demonstrated in Table 1, it is obvious that there are 220 positive samples in all subjects (Ct<38) and 200 negative samples (Ct>38). For the test results of antigen kit, the positive samples with the Ct⩽25 can be completed detected, which is totally in agreement with the real time PCR; For the positive samples with the Ct in the range from 25 to 30, 9 of 10 samples can be measured, indicating the detection percentage of 90%; For 30<Ct<33, the positive detection percentage is still as high as 86.6%; And the weak positive samples with the Ct in the range of 33<Ct<38, 3 positive samples were measured from the 7 samples with the positive detection percentage of 42.9%. For all negative samples (Ct>38) confirmed with real time PCR, no positive result was reported of antigen detection kit, indicating the 100% consistence with real time PCR.

**Table 1.** The comparison results between the colloidal gold SARS-CoV-2 antigen detection kit and the real time PCR according to the classification of different Ct values.

| **Ct values** | **SARS-CoV-2 antigen detection kit (n=420)** |  |  |
| --- | --- | --- | --- |
|  | **No. of Samples** | **Antigen positive** | **Detection percentage %** |
| **$\leq 25$** | 190 | 190 | 100 |
| **25-30** | 10 | 9 | 90 |
| **30-33** | 13 | 11 | 84.6 |
| **33-38** | 7 | 3 | 42.9 |
| **$> 38$** | 200 | 0 | 0 |

Furthermore, all the enrolled subjects were tested with our constructed method and the results were analyzed according to the categories of the ages. From the results shown in Table 2, it can be seen that positive samples in all age distributions can be well measured with the satisfied coincidence (97.27%) compared with the real time PCR confirmed positive results. These results indicate that this developed method can be applied for COVID-19 screening in all populations with different ages.

**Table 2.**
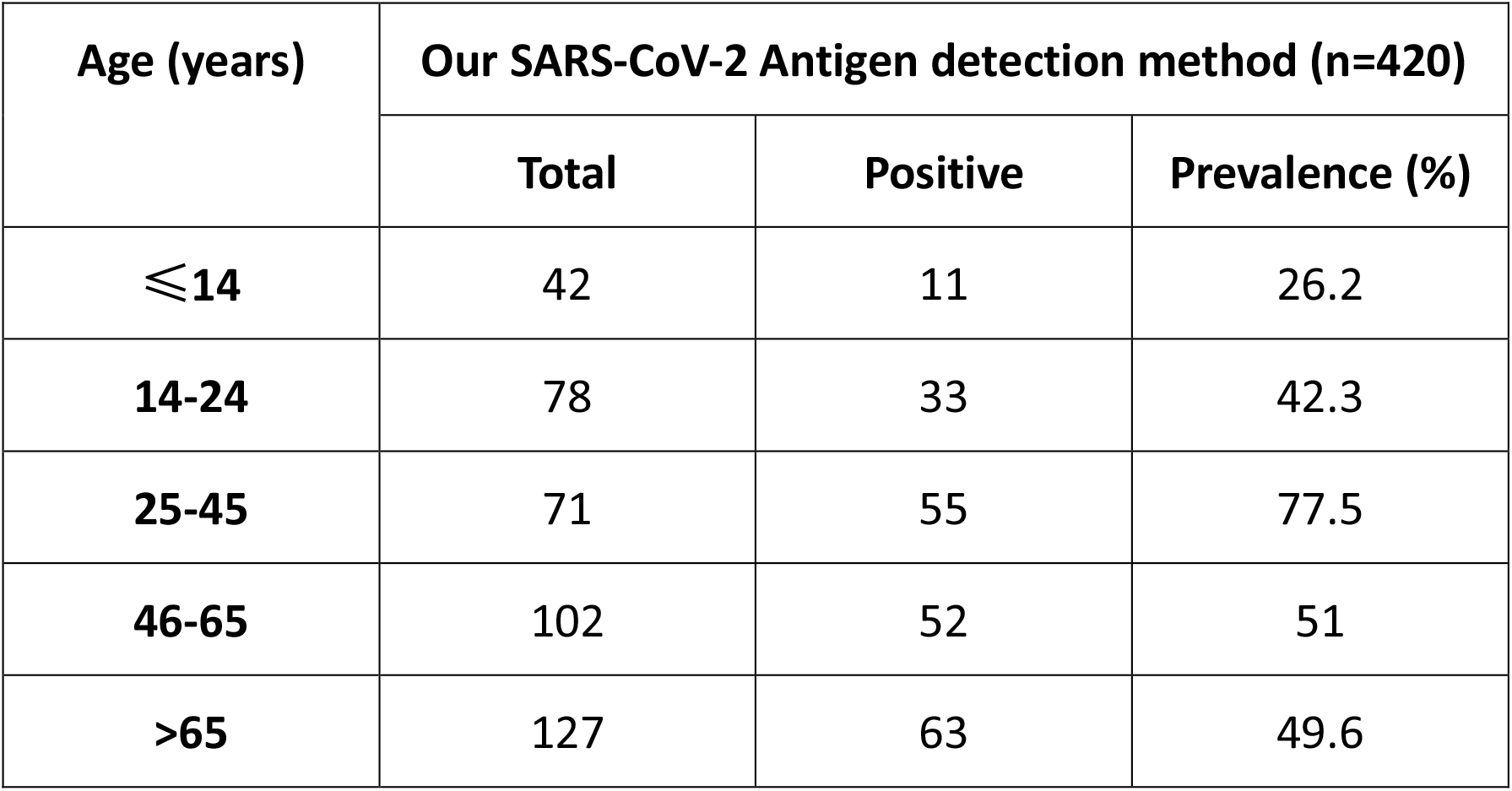
The antigen detection results of SARS-CoV-2 with colloidal gold kit (analyzed based on age distribution)

Besides, the detection results were further analyzed based on the classification of onset time of the symptoms. Detailed results expressed in Table 3 depicted the screening performance of the SARS-CoV-2 Antigen detection kit for samples with different onset time. In all enrolled subjects including both the positive and negative, the positive screening percentage is about 49.5% for the subjects with the onset time no more than 3 days while the positive screening percentage can be as high as 63.3% for the subjects with the onset time from 4 to 7 days. Considering the enrollment subjects, all these results suggest that the developed SARS-CoV-2 Antigen detection kit can meet the requirement of practical routine screening of large populations.

**Table 3.**
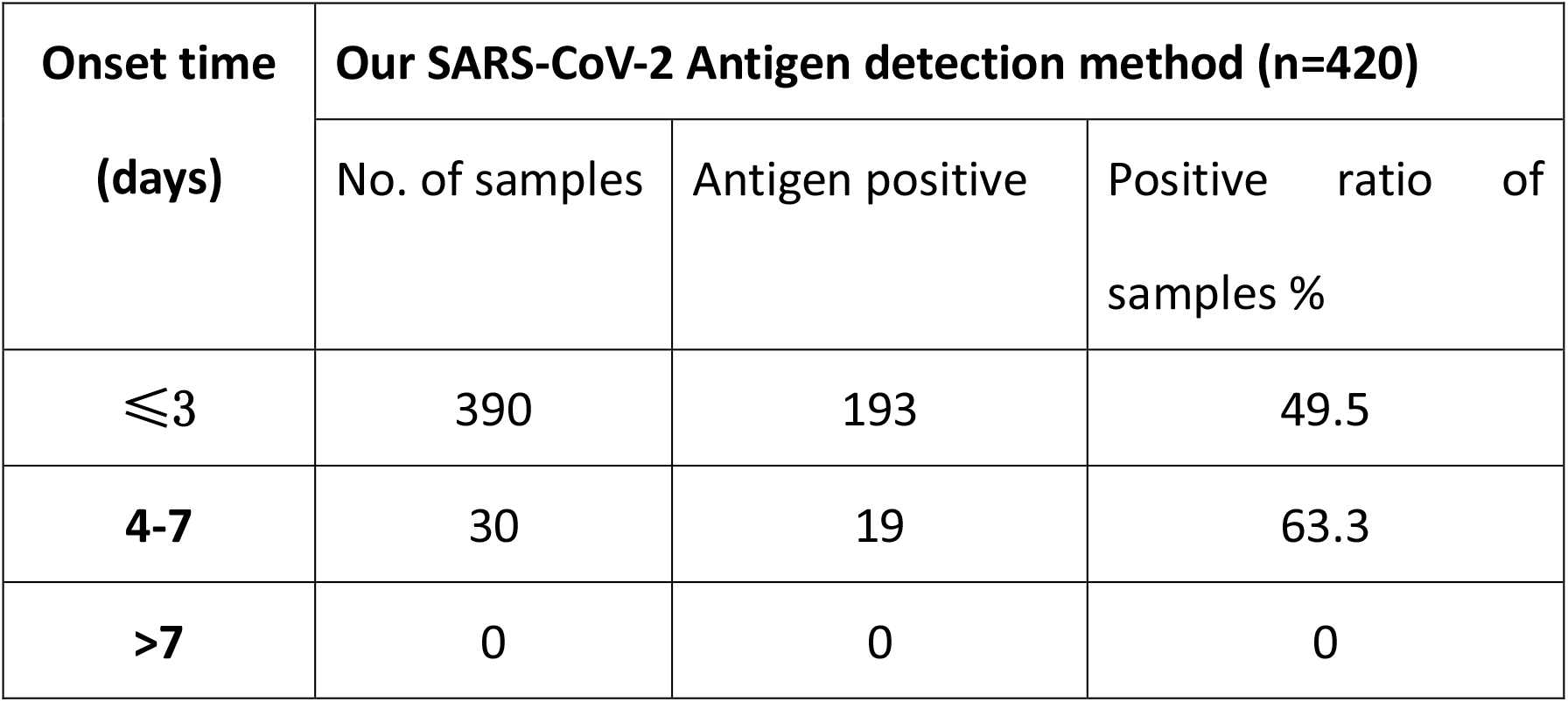
The screening results of SARS-CoV-2 Antigen detection kit for all subjects with different onset time of symptoms.

Finally, based on all above evaluation results, we have systematically analyzed the screening performance of this SARS-CoV-2 Antigen detection kit. The positive percentage agreement (PPA, sensitivity), negative percent agreement (NPA, specificity), the positive predictive value (PPV) and the negative predictive value (NPV) have been analyzed.

From the results shown in Table 4, the positive and negative percentage agreement of the SARS-CoV-2 Antigen detection kit are 96.8% and 98.1%, respectively compared with the real time PCR. Besides, for the routine screening of the large population, the PPV and NPV are 100% and 96.6%, respectively, which indicating the excellent potential for practical applications.

**Table 4.**
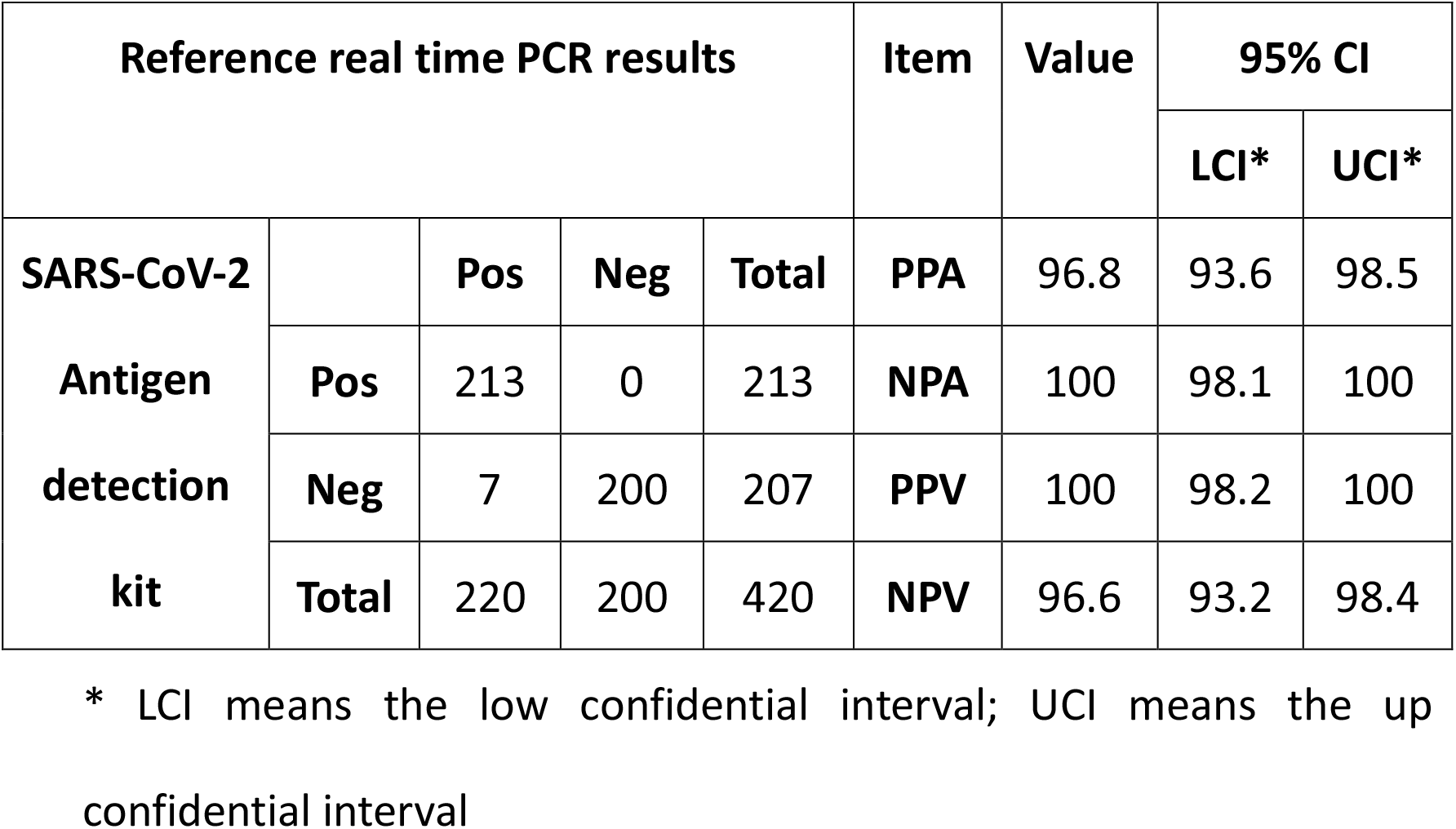
The systematic results analysis and performance evaluation of the SARS-CoV-2 Antigen detection kit.

Finally, the developed SARS-CoV-2 Antigen detection kit was also adopted for the screening of the delta variants of SARS-CoV-2. For the results shown in Figure 1, the developed SARS-CoV-2 Antigen detection kit can realize the successful detection of clinical delta samples. Of note, when the clinical samples were diluted with 10 or 100 times (correspond to the Ct values of 31 or 34), the SARS-CoV-2 Antigen detection kit could still demonstrate the positive results. Besides, the cultured delta virus strain sample (with the original Ct values of 22 or 23) was diluted for 16000 times and then measured directly with the SARS-CoV-2 Antigen detection kit. Results demonstrated that the positive signal was still very strong even after 16000-fold dilution of the sample. All these results well prove that this SARS-CoV-2 Antigen detection kit can be used for the practical screening of COVID-19 of the large population and pandemic control.

**Figure 1.**
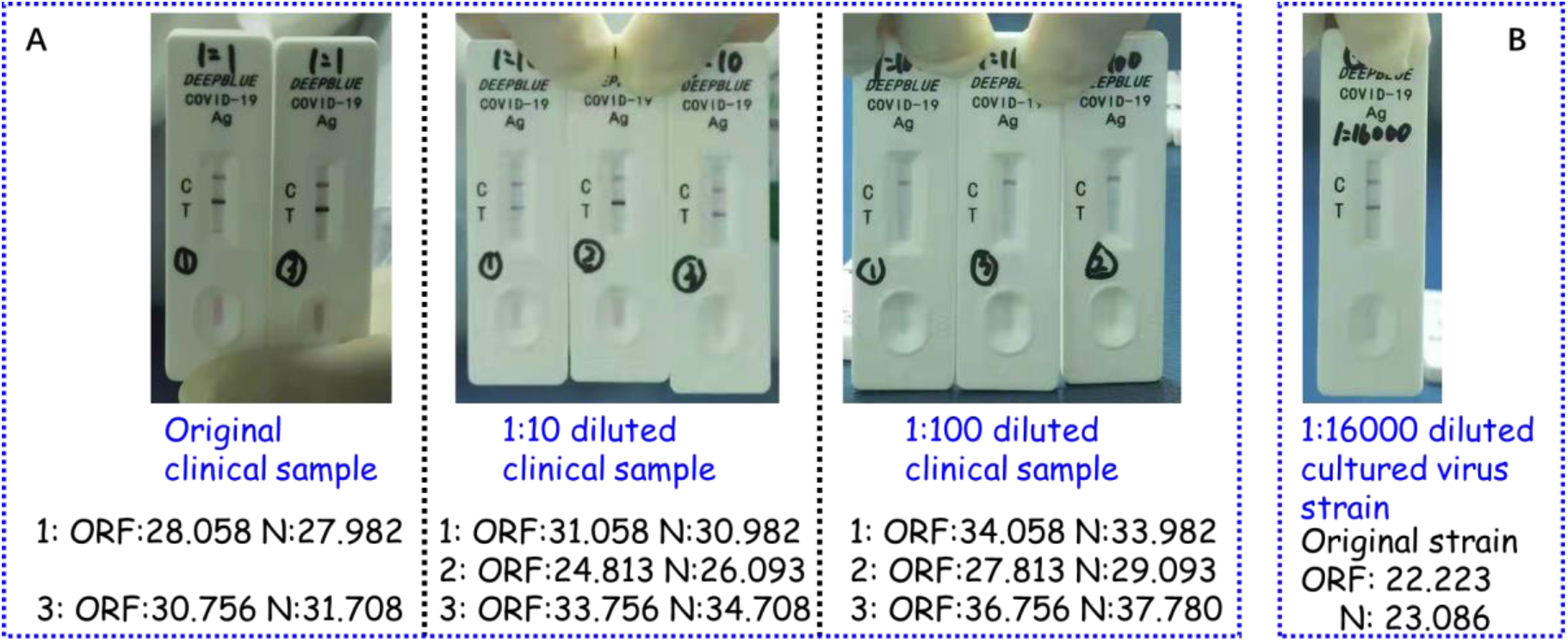
The test results of delta variant with the SARS-CoV-2 Antigen detection kit. (A. the test results of the clinical samples and diluted clinical samples; B. the tested results of the diluted cultured delta virus strain)

### Conclusion

In this research, we have developed and constructed the colloidal gold labeling principle based lateral flow strip for rapid and accurate detection of N protein of SARS-CoV-2. With the optimization of raw materials and detection conditions, this SARS-CoV-2 antigen detection kit has been used for detection of the clinical samples in the perspective model to verify the detection performance of the SARS-CoV-2 antigen detection kit. Results demonstrate that the detection results of the SARS-CoV-2 antigen detection kit are comparable to those of the real time PCR for the clinical samples from the enrolled subjects. The sensitivity and specificity of the SARS-CoV-2 antigen detection kit have been obtained as 96.8% and 98.1%, respectively. And the SARS-CoV-2 antigen detection kit has also been well applied in detection of delta variant of SARS-CoV-2. Satisfied results have also been achieved for delta variants detection, indicating the powerful potential for the routine screening of the large population of suspected objects.

## Data Availability

All data produced in the present study are available upon reasonable request to the authors.

## Acknowledgement

We would like to thank the Uniwersyteckie Centrum Kliniczne, Poland and the Anhui Provincial Center for Disease Control and Prevention, China for their professional verification of clinical samples in the professional BSL-3 labs.

